# Spatio-temporal modelling of referrals to outpatient respiratory clinics in the integrated care system of the Morecambe Bay area, England

**DOI:** 10.1101/2023.08.03.23293543

**Authors:** Rachael Mountain, Jo Knight, Kelly Heys, Emanuele Giorgi, Timothy Gatheral

**Affiliations:** Lancaster Medical School, Lancaster University, UK; University Hospitals of Morecambe Bay NHS Foundation Trust, Westmorland General Hospital, Kendal, UK

**Keywords:** Chronic respiratory disease, Integrated care, Outpatient referrals, Routinely collected data, Spatio-temporal

## Abstract

**Background:** Promoting integrated care is a key goal of the NHS Long Term Plan to improve population respiratory health, yet there is limited data-driven evidence of its effectiveness. The Morecambe Bay Respiratory Network is an integrated care initiative operating in the North-West of England since 2017. A key target area has been reducing referrals to outpatient respiratory clinics by upskilling primary care teams. This study aims to explore space-time patterns in referrals from general practice in the Morecambe Bay area to evaluate the impact of the initiative.

**Methods:** Data on referrals to outpatient clinics and chronic respiratory disease patient counts between 2012-2020 were obtained from the Morecambe Bay Community Data Warehouse, a large store of routinely collected healthcare data. For analysis, the data is aggregated by year and small area geography. The methodology comprises of two parts. The first explores the issues that can arise when using routinely collected primary care data for space-time analysis and applies spatio-temporal conditional autoregressive modelling to adjust for data complexities. The second part models the rate of outpatient referral via a Poisson generalised linear mixed model that adjusts for changes in demographic factors and number of respiratory disease patients.

**Results:** The first year of the Morecambe Bay Respiratory Network was not associated with a significant difference in referral rate. However, the second and third years saw significant reductions in areas that had received intervention, with full intervention associated with a 31.8% (95% CI 17.0-43.9) and 40.5% (95% CI 27.5-50.9) decrease in referral rate, respectively.

**Conclusions:** Routinely collected data can be used to robustly evaluate key outcome measures of integrated care. The results demonstrate that effective integrated care has real potential to ease the burden on respiratory outpatient services by reducing the need for an onward referral. This is of great relevance given the current pressure on outpatient services globally, particularly long waiting lists following the COVID-19 pandemic and the need for more innovative models of care.

## 1 Background

Chronic respiratory disease (CRD) remains a leading cause of morbidity and mortality in the UK; it is estimated that 15% of the population have a history of CRD and it is the fourth most common cause of death in England [1, 2]. Respiratory disease disproportionately affects disadvantaged socio-economic groups due to the known links with risk factors such as smoking, air pollution, poor housing, and occupational hazards [3]. CRD represents a large burden on the NHS with estimated direct costs of £4.7 billion from asthma and chronic obstructive pulmonary disease (COPD) alone [4]. The pressure is set to increase with an ageing population [5] which raises questions about how respiratory services can be changed to be more efficient and provide the best possible care for patients.

Promoting integrated care is a key goal of the NHS Long Term Plan to improve population respiratory health [6]. Integrated care is an organising principle for care delivery that seeks to improve the quality of care for patients by providing services that are better coordinated and act in a joined-up way [7, 8]. Integrated care has been argued as the key to making the health and social care system more sustainable. Without integration patients are more likely to become lost in the system, needed services can be duplicated or delayed, and the potential for cost-effectiveness declines [9]. However, despite the large push towards building integrated systems of care across England in recent years, evaluations have historically produced mixed results [10, 11, 12]. Research suggests this could, at least partly, be caused by the challenge in selecting outcome measures that are able to quantify the success of complex and multi-faceted initiatives [10, 13, 14]. The issue is exacerbated by data access challenges, particularly in primary and community care, that can be the limiting factor in possibilities for evaluations [14, 15].

The North-West region has the highest under 75 mortality rate from respiratory disease in England, 44.7% compared to 33.6% country-wide [16]. Clinical commissioning groups were dissolved on 1^st^ July 2022, but at the time of this analysis, they were NHS bodies responsible for the planning and commissioning of health care services for their local area in England. The Morecambe Bay Clinical Commissioning Group (MBCCG) in the North-West of England provided primary care for approximately 352,000 patients across 32 general practices (GPs) [17]. The majority of patients reside in Lancaster, South Lakeland, and Barrow-in-Furness, covering both rural and urban town areas, as well as a range of socio-economic levels including some of the most deprived communities in the country [18].

The Morecambe Bay Respiratory Network (MBRN) is an integrated care system operating in the Morecambe Bay area that aims to improve the quality and efficiency of healthcare delivery for patients with the four most prevalent CRDs in the UK: asthma, COPD, bronchiectasis, and interstitial lung disease (ILD). The first phase of the initiative began in 2017, reaching 50% of the MBCCG population through 8 practices in the network clustered predominately in the Lancaster and Barrow-in-Furness localities (Figure 1). A second phase in 2019 extended that reach to 65%, but this research focuses on phase one. The core components of the MBRN include an enhanced primary care team that has direct access to specialist investigation and is closely supported by secondary care expertise via monthly multidisciplinary meetings. The MBRN promotes effective communication and shared pathways across healthcare tiers to ensure that patients receive consistent information and to remove unnecessary or duplicate appointments. A key metric for the MBRN has been the measured impact on outpatient referrals reflecting the goals of improved service efficiency, bringing care closer to the patient, and avoiding unnecessary referrals that come at a high cost for outpatient services and increases wait time for all patients.

**Figure 1.**
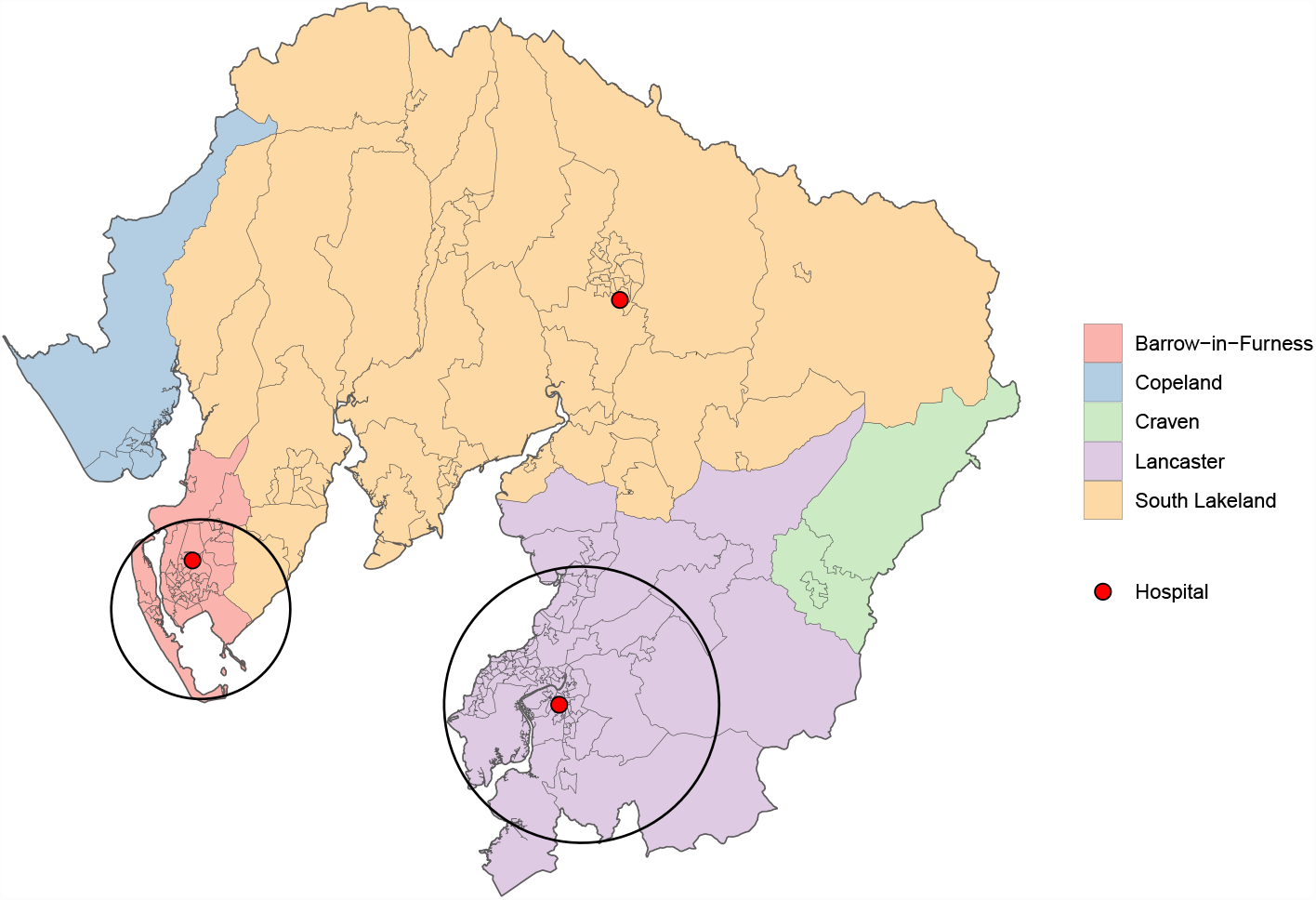
: Map of Morecambe Bay area shaded by local authority. Black circles show the approximate area of influence of the Morecambe Bay Respiratory Network.

The NHS undertakes 125 million outpatient appointments a year [19]. The COVID-19 pandemic has added considerable pressure to an already strained system with 6 million people on the waiting list for elective care compared to 4.4 million prior to the pandemic. The waiting list is expected to continue to grow in the short term as patients come forward who have delayed seeking health advice or treatment during the pandemic [20]. The radical redesign of elective care is more essential now than ever to manage demand in a way that improves patient care as well as service efficiency [21]. There is a need to work with general practice (GP) to improve patient pathways to reduce the need for an onward referral and avoidable delays where possible [20].

The aim of this research is to provide a data-driven assessment of the impact of the MBRN using a source of routinely collected data that has not been extensively used in health service research. The analysis will focus on an outcome measure of relevance to MBRN goals, referrals to outpatient respiratory clinics, whilst also adjusting for demographic factors and changes in CRD patient count to allow a closer study of the underlying referral behaviour. A novel aspect of our analysis is that we consider referral patterns at small area geography, unlike other studies that often analyse at individual or healthcare provider level. The remainder of the paper is structured as follows. After a brief overview of the modelling approach, we describe the routinely collected data source, including complexities and sources of missingness and the impact this may have on space-time analyses at small-area geography level. Next, we propose the methodology used that has two parts:

1. Spatio-temporal extension of conditional autoregressive models to adjust for the complexities in the data prior to the primary analysis.
2. Generalised linear mixed model of outpatient referrals in the Morecambe Bay area over an eight year period.

We then present the results of the model output before providing a concluding discussion, relating back to the impact of the MBRN, the wider context of the demand on outpatient services, and the importance of robust data for healthcare evaluations.

## 2 Methods

The main outcome variable is annual rate of referrals from GP to outpatient respiratory clinics over an eight year study period (1^st^ April 2012 -31^st^ March 2020) for 204 of the Lower-layer Super Output Areas (LSOAs) that lie within the MBCCG boundaries. LSOAs are small areas used for census geography in the UK that have an average population size of 1,500 [22]. The rate denominator of the outcome measure is number of diagnosed CRD patients to adjust for differences in patient count over space and time, and to avoid a model where referrals is acting as a proxy for prevalence. We consider data from adults aged 25 years or over. The 18-24 age bracket was excluded to reduce potential bias from the large student population in central Lancaster. Further, two LSOAs within the MBCCG boundaries were excluded due to the influence of Lancaster University.

For the sake of brevity, in the remainder of the paper study years will be referenced by the start date. For example, the study year ‘2012’ will refer to the period 1^st^ April 2012 – 31^st^ March 2013. Additionally, ‘adults’ will refer to individuals aged 25 years or over unless specified otherwise.

### 2.1 Primary data source

This study uses routinely collected NHS data stored in the Morecambe Bay Community Data Warehouse (CDW), a SQL Server owned and maintained by the University Hospitals of Morecambe Bay NHS Foundation Trust. The CDW contains data from primary, secondary, and community care across Morecambe Bay and uses pseudonymised NHS Numbers to allow linkage between data sets at an individual level.

Referrals were identified from secondary care records of the three hospitals within the study area with outpatient respiratory services: Furness General Hospital in Barrow-in-Furness, Royal Lancaster Infirmary in Lancaster, and Westmorland General Hospital in South Lakeland (Figure 1). A relevant referral was defined as any new referral from GP to a respiratory, spirometry, oxygen, or lung clinic, for an adult residing in the study area. We excluded referrals to clinics for asthma biologics, rheumatology, respiratory postoperative, respiratory physiotherapy, sleep apnea, and referrals made under the ‘two-week wait’ pathway for suspected respiratory cancer. The clinic inclusion-exclusion criteria were determined by physician expertise with the aim of capturing healthcare interactions that best aligned with MBRN goals and patient priorities.

Primary care records were used to build a GP-registered population dataset of all adults residing in a study LSOAs and registered at a MBCCG GP. An individual’s entry date is defined as the most-recent of GP registration start date and their 25^th^ birthday. Although registration status is recorded in the CDW, registration end date is missing for all individuals who have left or died so we use last interaction with primary care (appointment, consultation, or medication issue) as a proxy. An individual’s end date in the GP-registered population dataset, if relevant, is end date proxy or date of death.

CRD patients were identified from among the GP-registered population cohort by diagnoses recorded in primary care with a relevant asthma, COPD, bronchiectasis, or ILD SNOMED CT code. Relevant codes were identified using NHS Digital’s SNOMED CT Browser [23]. The codes were then filtered with the aim of reflecting as closely as possible MBRN’s own in-house patient registers. For an asthma diagnosis, an issuing of inhaled therapy in the past 12 months was used as an additional criterion. The Quality and Outcomes Framework guidelines [24] require post bronchodilator spirometry for a COPD diagnosis. We have not applied this criterion due to discrepancies in the recording of lung function test results in the CDW. A validation study found that using diagnoses codes alone gave a positive predictive value for true COPD of 86.5% and including spirometry results or medications only marginally improved results [25]. Start and end dates for diagnoses are recorded in the CDW and applied here to estimate the number of respiratory patients for any given space-time unit. In the case of asthma diagnoses, the ceasing of inhaled therapy for a period of 12 months qualifies as an end date.

The primary care data in the CDW has missingness and complexities that introduce bias to the GP-registered population cohort, in turn impacting the CRD patient counts. The three main issues are:

1. Two of the 32 MBCCG GPs are not signed up to the CDW data sharing agreement and so we do not have access to primary care records for these patients. This creates spatially-correlated gaps in the data.
2. We use a proxy for GP registration end date but this will likely be earlier than the true de-registration date, resulting in an underestimate of the GP-registered population size at any given time.
3. For a given registration, only a patient’s current address rather than entire address history is recorded and so movement of people within the MBCCG over time cannot be tracked. An individual’s current address is assumed to be the residency for their entire registration period which may result in individuals being assigned to an incorrect space-time unit.

Each of these issues has a spatial and/or temporal dimension and could bias the analysis via the denominator of the outcome rate.

### 2.2 Secondary data sources

GP registration data from NHS Digital [26] was used to estimate the GP-registered population counts that would be observed in the CDW without the presence of bias and missingness. Since 2014, NHS Digital releases data on a quarterly basis at LSOA-level for total number of patients registered at each GP practice in England. An age breakdown is not provided at LSOA level due to possible identification of individuals when linked to other data sets. However, an age breakdown is provided for each distinct GP register. Therefore, for each of the 204 LSOAs in the study area, we estimate the number of adults registered at a MBCCG GP by multiplying the number from the LSOAs population registered at each relevant GP by the proportion of that GP’s register ages 25 years or over, then summing across all GPs. This is repeated for all quarterly releases, and then averaged according to study year.

The Office for National Statistics (ONS) publishes mid-year population estimates that are used for estimates of LSOA age and sex demographics [27]. Although the census population and GP-registered population are not identical, we use the ONS estimates since NHS Digital does not cover all study years. The MBCCG and LSOA boundaries are also available from ONS as shapefiles [28, 29]. The Open Source Routing Machine (OSRM) package in R Studio was used to construct variables for distance to healthcare services [30]. The road distance in kilometres (km) was calculated for all postcodes within the study area and then averaged by LSOA.

### 2.3 Statistical Analysis

#### 2.3.1 Adjusting CRD patient count

We adjust the rate denominator, CRD patient count, for the previously described data complexities in the CDW GP registers by assuming that the population assigned to a given space-time unit by the CDW is representative of the corresponding true, unobserved GP-registered population. Then for study years 2014 onward, an estimate for the number of adult CRD patients is obtained for each LSOA by:

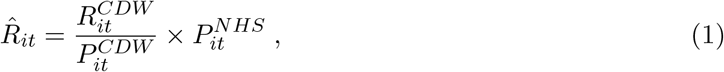

where 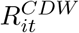 is the CRD patient count from the CDW for LSOA *i* (*i* = 1, …, *N*) and year *t* (*t* = 1, …, *T*), 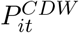 the GP-registered adult population count from the CDW, and 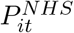 the GP-registered adult population estimate from NHS Digital. Since LSOA-level data is not available from NHS Digital pre-2014, we apply spatio-temporal modelling techniques to model the error in the primary care records of the CDW and to predict the NHS Digital figures for study years 2012 and 2013 based on the corresponding CDW count. Once the predictions are obtained, the adjustment in (1) can be applied.

The study period has *T* = 8 years (2012-2019), but for this model we also use data from 1^st^ April 2020 to 31^st^ March 2021 to improve prediction capacity. The outcome variable is 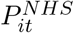 for LSOA *i* (*i* = 1, …, *N*) and year *t* (*t* = 1, …, *T* + 1). The counts are sufficiently large (mean = 1181, minimum = 681) to use a log-Gaussian model as an approximation to the Poisson. We include covariates for (natural logarithm of) CDW population count, 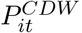, and measurable sources of error namely time and proportion of LSOA population registered at a GP not included in the CDW data sharing agreement (calculated using NHS Digital data). A generalised linear model (GLM) was first explored. The residuals exhibited strong spatio-temporal correlation: Moran’s I statistics computed on the residuals for each year separately produced values ranged from 0.23 to 0.33 with *p*-values less than 0.0001 in all years while the lag-1 temporal autocorrelation calculated for each LSOA separately yielded a mean of 0.3762 across all LSOAs.

We consider a model that captures the spatio-temporal autocorrelation via random effects assigned a spatio-temporal extension of conditional autoregressive (CAR) distributions, which are a type of Gaussian Markov random field. We assume the random effects to represent the unmeasured error in the CDW counts. Here we follow the model proposed by Rushworth et al. [31]. Let *S* = (*S*_1_, …, *S*_*T* +1_) denote the set of random effects for time points *t* = 1, …, *T* + 1, where *S*_*t*_ = (*S*_1*t*_, …, *S*_*Nt*_) is the vector of random effects for specific time point *t*. Then,

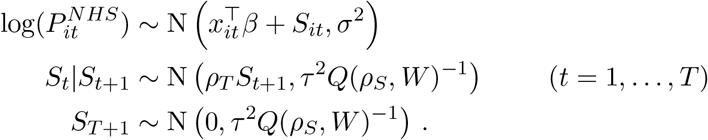

The vector *x*_*it*_ denotes the set of explanatory variables, *β* the corresponding regression parameters, and *σ*^2^ the variance of the residual errors. For the distributions of the random effects, *ρ*_*T*_ denotes the temporal dependency parameter, *ρ*_*S*_ the spatial dependency parameter, *τ*^2^ the conditional variance parameter, *W* an *N × N* neighbourhood matrix defined for the 204 non-overlapping spatial units that comprise the lattice data for this study, and *Q* the Leroux precision matrix [32]. Further detail for the spatio-temporal CAR model methodology can be found in the supplementary material.

The random effect for time point *T* + 1 is specified marginally since *S*_*T* +2_ is not observed. A typical first-order autoregressive process defines each value conditioned on the previous value. We condition in the reverse order since data is extracted from the CDW retrospectively making the most recent data the most accurate and error accumulating as we go further back in time.

#### 2.3.2 Modelling referrals to outpatient respiratory clinics

Let *Y*_*it*_ be the number of new referrals from GP to an outpatient respiratory clinic for LSOA *i* (*i* = 1, …, *N*) and year *t* (*t* = 1, …, *T*). The referral data is modelled using a Poisson generalised linear mixed model (GLMM) with a random intercept term for each LSOA, denoted by *Z*_*i*_. The adjusted number of CRD patients from the first part of the methodology, 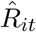, is included as an offset term to give a rate interpretation. Then,

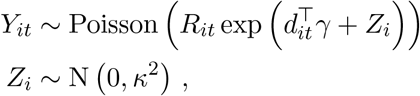

where *d*_*it*_ is the vector of explanatory variables, *γ* the corresponding regression parameters, and *κ*^2^ the variance of the random effects for which we assume independence. A corresponding Poisson GLM was over-dispersed yet exploratory analysis carried out on the residuals did not provide evidence to support a more complex correlation structure for the random effects (details given in the supplementary material file).

The covariate component of the GLMM is:

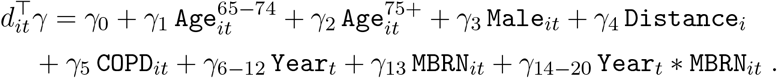

The covariates `Age`^65*−*74^, `Age`^75+^, and `Male` are included to account for demographic differences in the LSOAs and respectively represent the proportion of the adult population in the 65-74 and 75+ age brackets, and proportion of the adult population that are male. Stepwise covariate selection (with age groups 25-39, 40-54, 55-64, 65-74, 75+) suggested the age groups included are the only ones that are predictive of referrals and have a distinct effect to each other. Distance represents the average car travel distance to the nearest hospital within the MBCCG providing respiratory outpatient services.

The covariate `COPD` represents adult COPD prevalence and is primarily included to act as a proxy for socioeconomic deprivation, particularly that related to poor health [33]. The Index of Multiple Deprivation (IMD) [18] is the official measure of relative deprivation in England yet the index is only calculated every 3-4 years and considers a broad range of domains which may have less relevance to this research. Due to the correlation between IMD and COPD prevalence, we do not include both in the model and assume the latter to be a better indicator of deprivation related to the risk factors of respiratory disease.

To account for the MBRN effect at GP-level, we calculate the percentage of an LSOAs GP-registered population that is registered at a GP that joined the MBRN in 2017, represented in the model by `MBRN`. This is included for all years of the study period, even prior to MBRN introduction, to account for baseline differences in health service utilisation for LSOAs that received MBRN intervention from 2017 onward. Year represents the study year and `MBRN*Year` is an interaction term between study year and MBRN coverage, which will be the main indicator of the impact of the MBRN on outpatient referrals. Study year has been defined as a factor variable as opposed to a continuous covariate or a before/after MBRN indicator, in order to better study the evolution of MBRN impact since its initiation. For the sake of space, the factor levels have been grouped into one term in the above equation.

Additional descriptions of covariates used for both models can be found in the supplementary material file.

#### 2.3.3 Parameter Estimation

The models are specified as Bayesian hierarchical models and parameter estimation carried out using Markov Chain Monte Carlo (MCMC) algorithms. For the spatio-temporal GP registration model, the unobserved data for years 2012 and 2013 are treated as missing values in the response vector and are estimated each iteration via the posterior predictive distribution. When fitting the referrals model, to account for the uncertainty in the predictions, we randomly sample from the posterior samples for the predictions each iteration and recalculate the offset term. All statistical analysis was carried out in R Studio [34]. For further information on MCMC specifics, including prior distributions, we refer readers to the supplementary material file.

## 3 Results

### 3.1 Adjusting CRD patient count

Since the spatio-temporal model for patient count adjustment is not the main focus of this paper, we refer the interested reader to the supplementary material for extended results including covariate description, parameter estimates, prediction output, model validation, and MCMC diagnostics. Figure 2 and Table 1 are included here to highlight respectively the need and impact of the proposed adjustment modelling.

**Figure 2.**
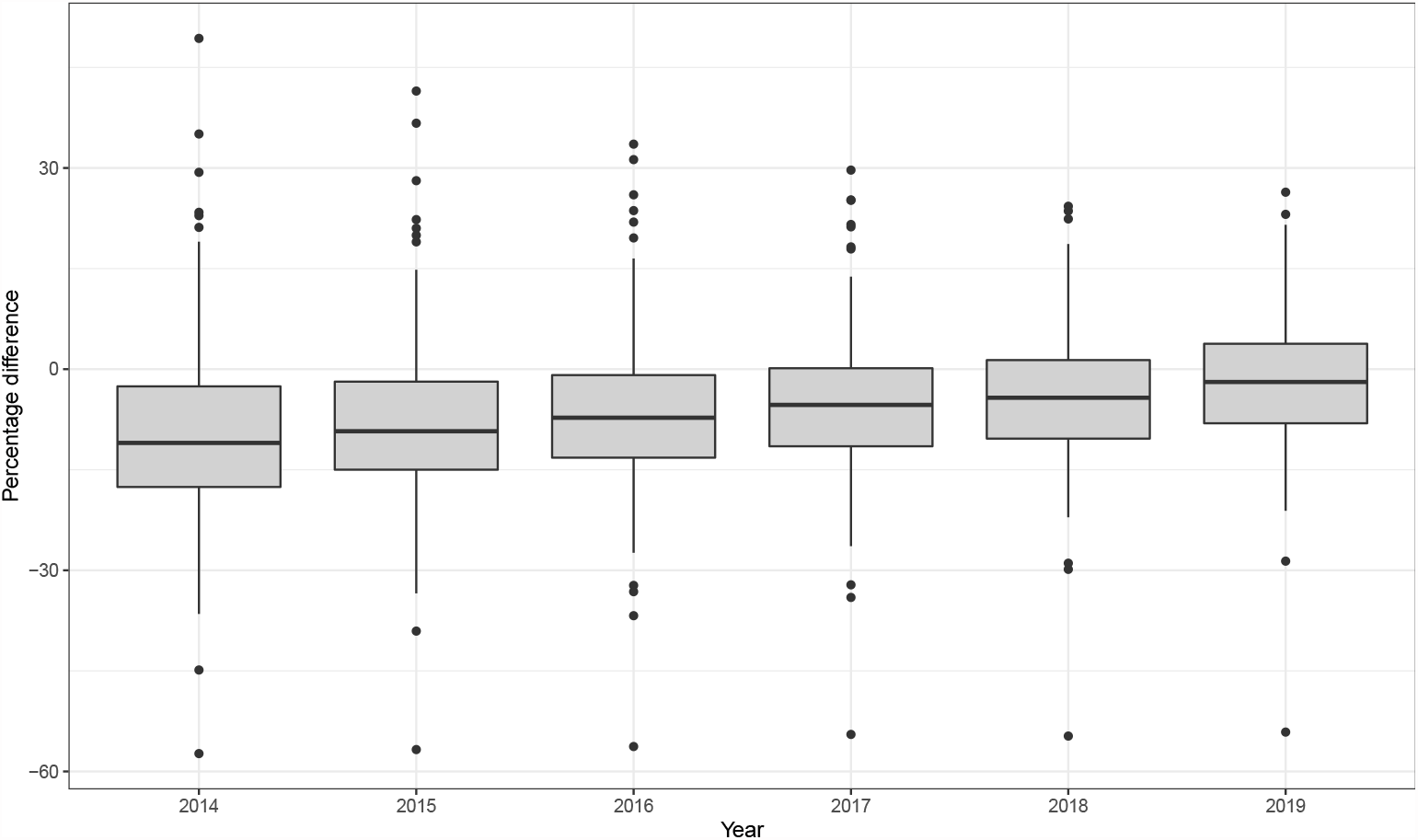
: Boxplot showing the spread of relative difference between the CDW counts and NHS Digital estimates for adults registered at a MBCCG GP at LSOA-level. Percentage difference = (CDW *−* NHS)*/*NHS *×* 100. Data for years 2012 and 2013 not available from NHS Digital.

**Table 1:**
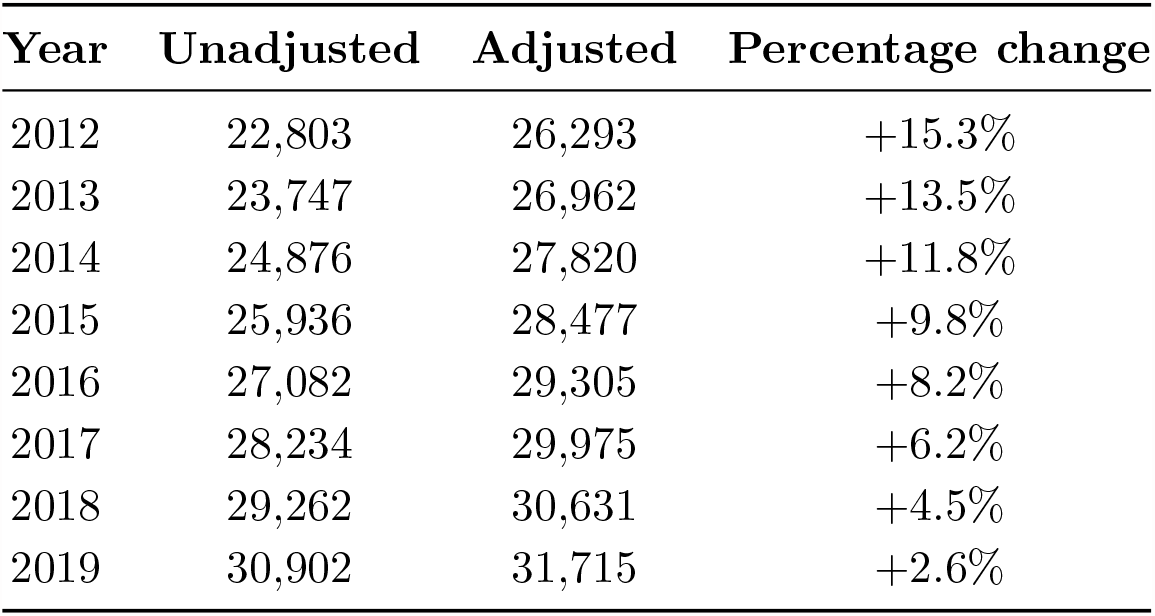
Comparison of unadjusted (raw counts extracted from CDW) and adjusted (prevalence *×* GP-registered adult population) total CRD patients within the MBCCG for each study year.

| Year | Unadjusted | Adjusted | Percentage change |
| --- | --- | --- | --- |
| 2012 | 22,803 | 26,293 | +15.3% |
| 2013 | 23,747 | 26,962 | +13.5% |
| 2014 | 24,876 | 27,820 | +11.8% |
| 2015 | 25,936 | 28,477 | +9.8% |
| 2016 | 27,082 | 29,305 | +8.2% |
| 2017 | 28,234 | 29,975 | +6.2% |
| 2018 | 29,262 | 30,631 | +4.5% |
| 2019 | 30,902 | 31,715 | +2.6% |

Figure 2 shows the spread of percentage change between the CDW GP-registered population counts and NHS Digital estimates for study years 2014-2019. As we go further back in time, the magnitude of the median percentage difference increases and there is increased variation in the degree of error. The plot illustrates the consequence of the issues described with the CDW GP registers. For example, the point that is consistently a 50-60% underestimate represents an LSOA where a large proportion of the population is registered at a GP not in the CDW data sharing agreement. The points around a 30% overestimate and above in 2014-2016 represent LSOAs that have undergone significant housing development. If individuals move into the houses from the local area, the CDW does not store the address history, thus assigning them to an address at a time before the housing existed.

Table 1 summarises the overall impact of the adjustment methodology on the total number of CRD patient counts used as the denominator of the outpatient referral rate.

### 3.2 Modelling referrals to outpatient respiratory clinics

#### 3.2.1 Raw data

We first present data summary results relating to the reach of the MBRN. The MBRN covariate in the GLMM is defined as a percentage of the total population, yet since the GPs in the network are clustered in the most urban areas of MBCCG (Figure 1), 84% of the data points are either less than 1% or greater than 99%, with an overall median of 51.4%. Table 2 shows a comparison of age, sex, distance to nearest hospital, and COPD prevalence for LSOAs that received MBRN intervention in 2017 and those that did not. For the sake of Table 2 we dichotomise the variable so that an LSOA is classed as ‘MBRN’ if `MBRN`*>* 50% and ‘Non-MBRN’ otherwise. From Table 2 it can be seen that the populations of the MBRN areas are on average younger, closer in distance to a major hospital, and have higher rates of COPD.

**Table 2:** Comparison of the median (IQR) of demographic covariates used in the GLMM between intervention and non-intervention areas. The MBRN covariate has been dichotomised at the 50% mark.

|  | **MBRN** | **Non-MBRN** |
| --- | --- | --- |
| 65-74 | 15.4 (12.1, 19.3) | 18.5 (14.7, 21.0) |
| 75+ | 12.6 (9.1, 16.6) | 14.0 (11.3, 17.0) |
| Male | 47.7 (46.5, 49.0) | 48.1 (46.8, 49.6) |
| Distance | 6.0 (2.4, 8.2) | 11.0 (3.76, 20.4) |
| COPD | 3.3 (2.5, 4.7) | 2.7 (2.0, 3.7) |

A total of 8,897 referrals to outpatient respiratory clinics that fulfilled the inclusion criteria were extracted from secondary care records in the CDW. Table 3 documents the raw counts by study year and the average number of referrals per LSOA. The total number of new referrals from GP to respiratory outpatient clinics displayed a consistent increasing trend up to 2016, but the counts in the years since the introduction of the MBRN (2017-2019) have not risen above 2016 levels.

**Table 3:**
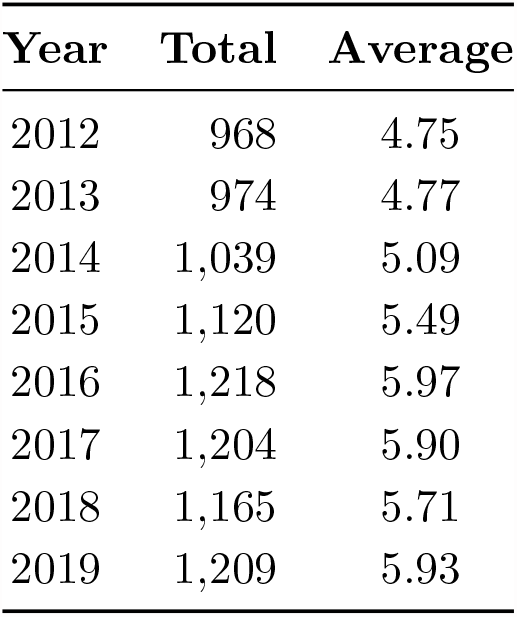
Total number and average per LSOA of new referrals to outpatient respiratory clinics from GP for each study year.

Additional data summaries can be found in the supplementary material.

#### 3.2.2 Model output

Table 4 presents the parameter estimates for the GLMM. The continuous covariates in the model represent percentages (e.g. percentage of LSOA population registered at an MBRN GP or percentage of LSOA adult population that are male), with the exception of the distance to nearest hospital covariate which is in kilometres. The relative risk interpretation is the risk increase for each unit (i.e. one percentage or one kilometre) increase in the covariate. As a result, the relative risks are of small magnitude with credible interval boundaries that can round to 1.000 to three decimal places. Therefore, the relative risk and 95% CIs associated with a one-standard deviation increase in the variable is also given to give a more meaningful interpretation in the context of the data.

**Table 4:** A summary of the parameter estimates from the outpatient referrals random intercept model. The median relative risks (RR) and 95% credible intervals (CI) are presented for the standard results and for a one standard deviation (std. dev) increase in each continuous variables, shown in the final column.

| Parameter | One unit increase |  | One std. dev increase |  |  |
| --- | --- | --- | --- | --- | --- |
|  | RR | 95% | RR | 95% | Std. dev |
| Intercept | 0.032 | (0.014, 0.074) | - |  |  |
| Age 65-74 | 1.016 | (1.007, 1.025) | 1.078 | (1.034, 1.125) | 4.70 |
| Age 75+ | 1.011 | (1.003, 1.018) | 1.061 | (1.014, 1.106) | 5.61 |
| Male | 1.012 | (0.999, 1.025) | 1.033 | (0.997, 1.070) | 2.75 |
| Distance to hospital | 1.005 | (1.001, 1.009) | 1.042 | (1.007, 1.079) | 8.47 |
| COPD | 0.962 | (0.944, 0.981) | 0.939 | (0.910, 0.969) | 1.64 |
| 2013 | 0.967 | (0.885, 1.058) |  | - |  |
| 2014 | 0.996 | (0.912, 1.087) |  | - |  |
| 2015 | 1.041 | (0.955, 1.135) |  | - |  |
| 2016 | 1.102 | (1.014, 1.199) |  | - |  |
| 2017 | 1.063 | (0.977, 1.158) |  | - |  |
| 2018 | 0.987 | (0.906, 1.074) |  | - |  |
| 2019 | 0.971 | (0.891, 1.059) |  | - |  |
| MBRN (*main effect*) | 1.002 | (1.000, 1.003) | 1.075 | (1.005, 1.149) | 42.79 |
| MBRN 2013 | 0.998 | (0.996, 1.000) | 0.918 | (0.841, 1.001) | 42.79 |
| MBRN 2014 | 0.999 | (0.997, 1.001) | 0.942 | (0.866, 1.025) | 42.79 |
| MBRN 2015 | 0.998 | (0.996, 1.000) | 0.913 | (0.840, 0.991) | 42.79 |
| MBRN 2016 | 0.999 | (0.997, 1.001) | 0.969 | (0.892, 1.053) | 42.79 |
| MBRN 2017 | 0.999 | (0.997, 1.001) | 0.958 | (0.882, 1.041) | 42.79 |
| MBRN 2018 | 0.996 | (0.994, 0.998) | 0.849 | (0.781, 0.923) | 42.79 |
| MBRN 2019 | 0.995 | (0.993, 0.997) | 0.801 | (0.738, 0.871) | 42.79 |

The main indication of the effect of the MBRN are the interaction terms between MBRN coverage and year. We first consider the interaction terms for the years prior to MBRN introduction (pre-2017). Since year is specified as a factor covariate, the MBRN main effect represents the interaction between MBRN and study year 2012. In 2012, a one-standard deviation increase was significantly associated with a 7.5% increase in referral rate and in 2015, a one-standard deviation increase was significantly associated with an 8.7% decrease in referral rate compared to 2012. However, note that the credible intervals for 2012 and 2015 are close to one even for the relative risk associated with a one-standard deviation increase. The years 2013, 2014, and 2016 are insignificant. The pre-MBRN interaction terms do not suggest a systematic difference in referral rates, after adjusting for all other covariates, for LSOAs that received higher percentages of MBRN intervention from 2017 onward.

The MBRN did not have a significant association with referral rate in the activation year (2017). In 2018, a 42.79% increase in percentage registered at an MBRN GP was associated with a 15.1% decrease in the rate of new referrals from GP. In 2019, the same registration percentage increase is associated with a 19.9% decrease in referral rate. To compare no MBRN intervention with full MBRN intervention, in 2018 and 2019 a 100% increase in MBRN coverage (i.e. from no coverage to full coverage) is associated with a 31.8% (95% CI 17.0-43.9) and 40.5% (95% CI 27.5-50.9) decrease in referrals, respectively.

The model output does not suggest a significant change in overall referral rate over time beyond what can be attributed to changes in demographic factors and the introduction of the MBRN. All levels of the year factor variable are insignificant except for 2016 which shows a significant 10.2% increase in referral rates compared to 2012.

Figure 3 provides a clearer illustration of the effect of MBRN coverage over time. The rate of referrals is predicted using the fitted model output for each time point and for three different levels of population registration at an MBRN GP (0%, 50%, and 100%). All other covariates including the offset term are held at their median values across the entire data set. In the baseline years (2012-2016) and in the activation year (2017), the credible intervals for the three predictions consistently overlap. In 2018, the prediction intervals begin to separate, with 0% continuing in an upward trend and 100% significantly decreasing, such that the two intervals do not overlap. In 2019 the separation widens, and an LSOA with just 50% registration has a credible interval entirely lower than that associated with 0% registration. In 2019, the model predicts a median rate of 3 referrals per 100 CRD patients (95% CI 2.7-3.4) for 100% MBRN registration compared to 4.3 per 100 CRD patients (95% CI 3.9-4.7) for 0% registration. Although the difference in rate may first appear small, applied to the MBCCG population that had an estimated 31,715 adults with a CRD diagnosis in 2019 (Table 1), the median reduction in rate equates to a difference of over 400 referrals a year: 951 referrals under full MBRN intervention compared to 1,364 under no MBRN intervention. It can also be seen from Table 4 that of the other covariates in the model, all are significant except proportion male. The 65-74 bracket has the more substantial effect of the two age covariates, although both display a positive association with the outcome measure. For each 4.7% increase in the proportion of an LSOAs adult population in the 65-74 age bracket, a 7.8% increase in referral rate would be expected. For the 75+ age bracket, the corresponding figures are 5.61% and 6.1% respectively. Distance to closest hospital is also positively associated with increased referral rate with an increase of 4.2% associated with an 8.47 km increase in road travel distance. For a 1.64% increase in an LSOAs COPD prevalence, a decrease in referral rate of 6.1% would be expected, suggesting a negative relationship between rate of referral and socio-economic deprivation.

**Figure 3.**
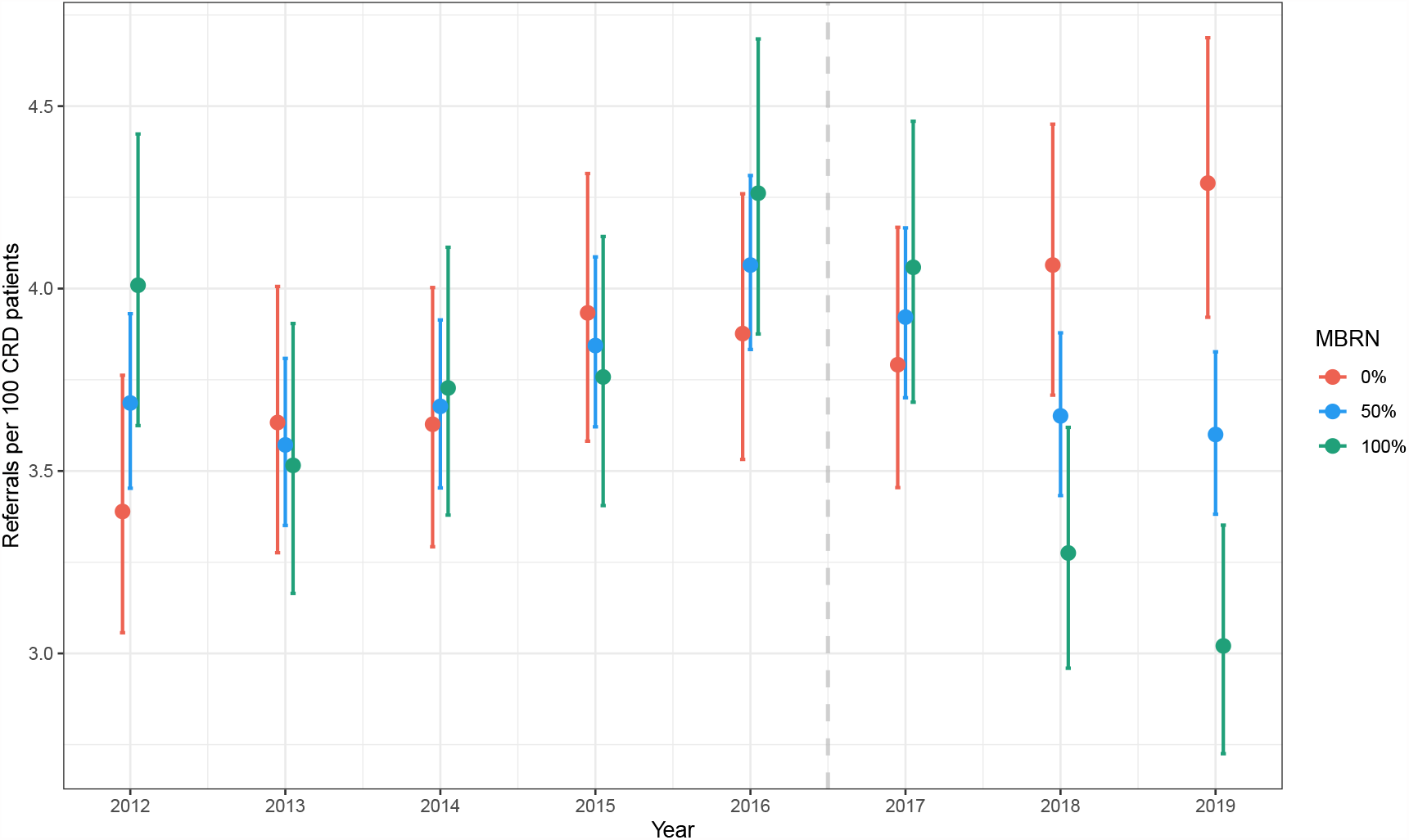
: The effect of the interaction term between Year and proportion registered at an MBRN GP on referral rate over time. The predicted number of referrals per 100 CRD patients is estimated for three levels of registration at an MBRN GP at LSOA-level. The dashed line represents the introduction of the MBRN in 2017.

The variance term for the random effects, *κ*^2^, was estimated at 0.021, 95% CI (0.014, 0.032), supporting the need for a GLMM over a GLM.

Diagnostics for the MCMC algorithm can be found in the supplementary material.

## 4 Discussion

Integrated care is a broad concept with multi-faceted implications which presents a challenge for evaluators. This study considered the use of routinely collected data to provide a robust data-driven analysis of healthcare delivery that focuses on an outcome measure of relevance and adjusts for diversity in the study population. The results suggest the success of the MBRN model in reducing rates of new referrals to outpatient respiratory clinics from GP in areas that have received higher percentages of intervention compared to areas with lower intervention. The effect of the MBRN progressively increased between 2017-2019 highlighting the importance of evaluating healthcare initiatives over sufficient time periods [14, 35]. Policymakers frequently want to see immediate results yet transformational changes in practice and work culture requires time to gain traction [36].

The first stage of the methodology in this paper applies existing saptio-temporal methodology to a new setting to model official statistics and predict beyond the published time frame at the required geography level based on error-filled routinely collected data. In addition, we account for uncertainty in the predictions by using the full posterior predictive distributions in the model fit for the mixed Poisson model. The methodology could be used in other fields of research that use time restricted official statistics such as further areas of health service provision, public health and social care, and the broader social sciences.

This research supports existing literature that integrated care services for a specific chronic disease using an MDT approach with disease-specific specialist input is likely to be successful [11, 37]. The MBRN shows the potential for effective integrated care with a broader patient scope and more ambitious vision for transforming CRD healthcare delivery. This provides new insight as existing integrated services for respiratory disease commonly focus on COPD alone, and often target only the most high-risk patients to prevent non-elective hospital admissions [38, 39, 40].

The results identified a negative relationship between COPD prevalence and rate of referrals. If COPD prevalence is effectively acting as the intended proxy for socio-economic position and general ill-health, then this finding supports existing literature that the most disadvantaged often have lower probabilities of attending specialist care [41, 42]. This study is unable to comment on the reason for the inequality in the MBCCG context; possible reasons include patient preference [41], lack of adequate communication [43] or health literacy [44], or differences in GP referral behaviour across the study area [45]. Existing research predominately does not find an association between socio-economic position and probability of visiting primary-care in developed countries, with some studies even reporting higher rates of attendance [41, 46]. There is an opportunity for integrated care services to reduce healthcare inequalities by training primary care to provide more specialist services.

Of the remaining covariates in the model, the 65-74 age bracket was at the greatest risk of higher rates of referrals whilst sex did not have a significant relationship. Distance to hospital was positively associated with rate of referral, which contrasts with existing research that links rurality with reduced access to services [47]. A probable explanation for this finding is the relative affluence of the rural areas in the MBCCG. We are unable to identify previous work in which outpatient referrals have been analysed at an LSOA-level in England. The study area contained 32 GPs but 204 LSOAs, hence using the small-area geography gave potential for insight into the contribution of other risk factors for rate of referral that may have been lost if covariates were averaged to GP-level. This is a particular issue in the MBCCG where several practises are made up of multiple sites. For example, Lancaster Medical Practice is comprised of eight separate sites spread over central Lancaster, serving a wide spectrum of patients, demographically and clinically speaking.

The recovery of elective care following the COVID-19 pandemic is not unique to the NHS but is affecting healthcare systems worldwide [48]. The NHS post-COVID recovery plan states the need for an increase in activity of 30% above pre-pandemic level by 2024/25 to reduce waiting times, but this goal has been met with scepticism in light of NHS staff shortages and recruitment challenges [20, 49, 50]. The MBRN model demonstrates that effective integrated care has real potential to ease the burden on respiratory outpatient services by reducing the need for an onward referral through improved patient pathways, effective communication between healthcare tiers, and an upskilled primary care team. Assuming a respiratory clinician has 2-3 4-hour clinics per week and assigns 30 minutes to a new patient [51], the reduction of 400 new referrals per year in the MBCCG population if all GPs were connected to the MBRN would equate to approximately one fewer clinic per week, with no consideration made for the knock-on effect to follow-up appointments.

A key strength of the proposed methods is using number of diagnosed CRD patients as the referral rate denominator. Disease prevalence data is not always readily available particularly at small area geography level and changes in patient counts, beyond what is able to be accounted for through population growth and known risk factors, can distort both space and time analyses of healthcare utilisation [52, 53]. In addition, integrated care initiatives often target improved diagnostic capacity and identify unmet need in their populations, resulting in evaluations reporting an increase in total healthcare service usage [54]. The model in this paper controls for changes in the size of the patient cohort, allowing a closer study of the underlying referral behaviour.

Data is vital for understanding the impacts of health interventions and generating robust analytics to improve healthcare delivery [35, 55]. The level of detail available in the CDW facilitated the flexibility of this analysis, including choice of outcome measure, spatial unit, adjusting for changes in CRD patient size, and filtering referrals and patients at a finer scale to capture healthcare interactions of closest relevance to the MBRN. This research is the first extensive use of the CDW for health service research and has barely scratched the surface of its potential. The CDW uses pseudonymised NHS Numbers; linking data at a patient level and removing traditional data silos between different branches of healthcare has the potential to provide a far more realistic and holistic view of patient care pathways. There is a clear, high value in investing in databases and personnel to exploit the wealth of information available in routinely collected data to support evidence-based decision making [56, 57].

The limitations associated with routinely collected data are well-established [58, 59]. This research contributes to the existing literature by exploring limitations encountered when using primary care records for space-time analyses, particularly the difficulty in tracking movement of people. The methodology proposed to circumvent the issues identified is somewhat of a crude fix and relies on the assumption that the prevalence calculated from the CDW is representative of the true, unobserved adult population for the corresponding space-time unit. This assumption may not be reasonable for error introduced by movement of people due to the relationship between transiency and age. If an age breakdown was provided at LSOA-level by NHS Digital, then a more informed adjustment to CRD patient count could be considered. Nevertheless, the strong spatio-temporal correlation identified in the CDW error process may be useful for future research into methodology for improving analysis using routinely collected healthcare data.

Other limitations of this research must be recognised. First, this is not a controlled study. The data summary results evidence that the MBRN reaches the most urbanised and deprived areas of the MBCCG. However, exploratory analysis (found in the supplementary material) and the interaction terms in the GLMM prior to MBRN introduction did not suggest a systematic difference in referral rates, once all else adjusted for, between intervention and non-intervention areas. Therefore, it is reasonable to attribute the dramatic decrease in referrals in 2018 and 2019 to the work of the MBRN. However, areas that have not received MBRN intervention may not have the same capacity for referral reduction due to potential differences in disease severity and patient need, which are not accounted for in the model. Second, in the GLMM, year is defined as a factor variable, adding to model complexity and forcing the relative risks to be compared to a baseline year, in this case 2012. The factor form was used over other representations, such as a linear time trend or a before/after indicator variable, as it best captured the evolving impact of the MBRN between 2017-2019. Due to the small temporal sample size, more advance time series modelling was not appropriate. Third, the study period was restricted by the introduction of the first COVID-19 lockdown in England, so we cannot comment on whether the reduction in referral rate was sustained. Finally, reason for referral is not documented in the secondary care records in the CDW therefore it is likely that the referrals modelled in this paper include irrelevant referrals despite efforts made with clinic inclusion-exclusion criteria.

This research evaluates the value of an integrated care initiative from a service utilisation point of view, yet it is unable to offer insight into the benefits on individual patient standard of care. This paper is the first in a series of research that utilises the CDW to evaluate the impact of the MBRN on respiratory healthcare delivery in the Morecambe Bay area. Future research will continue to focus on indicators that reflect the specific goals of the MBRN whilst homing into individual care patterns and clinical benefits, namely diagnostic quality and symptom control following diagnosis.

### 4.1 Conclusion

Overall, our novel analysis demonstrates the use of large routinely collected data to robustly evaluate key outcome measures of integrated care, in this case, rate of referrals to outpatient services. The results of this study is of great relevance in the current economic climate given the demand for innovative models of care and the large outpatient service backlog, a pressure felt by healthcare services across the globe. Future work should focus on assessing appropriate measures of quality for diagnosis in primary care and patient outcomes.

## Data Availability

The primary data of this study is routinely collected NHS data provided by the University Hospitals Morecambe Bay NHS Foundation Trust. The data cannot be shared publicly. However, other socio-economic and demographic data are publicly available. Website links for raw data extract can be found in the references. The aggregated forms used for this analysis can be made available from the corresponding author on request.

## Acknowledgements

We thank the Data Science unit at Royal Lancaster Infirmary for their assistance in constructing the data sets used for this research. We also thank the Morecambe Bay Respiratory Network for their insight into respiratory care and input on project design.

## Funding

RM is funded by an Economic and Social Research Council, which is part of UK Research and Innovation, doctoral training partnership (grant number: ES/P000665/1). JK is supported by Health Data Research UK, which is funded by the UK Medical Research Council, Engineering and Physical Sciences Research Council, Economic and Social Research Council, Department of Health and Social Care (England), Chief Scientist Office of the Scottish Government Health and Social Care Directorates, Health and Social Care Research and Development Division (Welsh Government), Public Health Agency (Northern Ireland), British Heart Foundation and the Wellcome Trust

### Abbreviations

CAR: conditional auto-regressive.
CDW: Community Data Warehouse.
COPD: chronic obstructive pulmonary disease.
CRD: chronic respiratory disease.
GLLM: generalised linear mixed model.
GLM: generalised linear model.
GP: general practice.
ILD: interstitial lung disease.
IMD: Index of Multiple Deprivation.
Km: kilometres.
LSOA: Lower-layer Super Output Area.
MBCCG: Morecambe Bay Clinical Commissioning Group.
MCMC: Markov chain Monte Carlo.
MBRN: Morecambe Bay Respiratory Network.
NHS: National Health Service.
ONS: Office for National Statistics.

## Ethics approval

All methods were carried out in accordance with relevant guidelines and regulations. This research has received ethical approval through the ethical committees of Health Research Authority and Health and Care Research Wales (IRAS project ID: 289188). The primary data of this study is anonymised routinely collected NHS data so the ethical committees of Health Research Authority and Health and Care Research Wales waived the need for informed consent from subjects to participate.

## Authors’ contributions

RM engaged in project design, processed the data, carried out statistical analysis, wrote the article, prepared all figures and tables, and was responsible for final submission of the manuscript. JK engaged in project design, project management, and was involved in editing the manuscript. KH engaged in project design, data curation, and data consultation. EG engaged in project design and statistical development, and was involved in editing the manuscript. TG was engaged in project design, provided clinical interpretation and application, and was involved in editing the manuscript. All authors approved the final manuscript.

