## Supplementary material for "Spatio-temporal modelling of referrals to outpatient respiratory clinics in the integrated care system of the Morecambe Bay area, England"

#### 1 Data

##### 1.1 Primary data source - additional description

The construction of the GP-registered population dataset from the CDW is outlined in the main text. We made the following additional choices:

- Duplicate NHS Numbers (most commonly caused by an individual being registered at more than one GP) with agreeing sex, date of birth, and date of death (if applicable), are assumed to be the same person, and the record with the most recent GP registration start date is taken as their current GP practice and address. If there is a disagreement in one of the aforementioned variables, then all records with the given NHS number are excluded.
- In order to be counted in a particular year, an individual's entry date must be prior to the half way point (1st October) of the given year and their end date after the halfway point. This was done to avoid overestimating the population count, particularly in areas with highly transient populations.
- For a given GP registration in the CDW, only the individual's current address, rather than entire address history, is recorded. A change in address can only be identified if it is accompanied by a change in registered GP. Consequently, it is possible to observe "large" moves in people, but not "smaller" local moves. This limitation is exacerbated by several GP practices in the MBCCG being made up of multiple sites. For example, Lancaster Medical Practice is comprised of eight separate sites spread over central Lancaster, hence an individual could move multiple times living in varied areas, demographically speaking, whilst remaining with the same GP. In addition, since GP registration end date is missing, it is not possible to determine whether there are breaks between registrations, for example if an individual has moved out of the MBCCG then moved back in at a later date. Therefore, we only consider the most recent registration for each distinct individual. Although this does waste some information,

given the relatively short length of the study period it should only impact majorly on areas with transient populations which will have a spatial correlation.

- 'Regular' registrations only are considered. In England, a 'temporary' GP-registration can be used while away from home for work, study or on holiday for up to 3 months. Individuals with a temporary registration remain registered with their permanent GP surgery during this time.

#### 1.2 Covariate description

Tables S1 and S2 provide a summary of the covariates used in the two models, including data source, general description, and additional notes.

Table S1: Description of variables used in spatio-temporal GP registration prediction model

| **Variable** | **Time varying (Y/N)** | **Source** | **Description** | **Notes** |
|---|---|---|---|---|
| GP-registered population | Y | NHS Digital | Average number of adults (25+ years) registered at a MBCCG GP for each LSOA inside the CCG boundaries | LSOA-level data released quarterly ($1^{st}$ Jan, $1^{st}$ Apr, $1^{st}$ Jul, $1^{st}$ Oct) since January 2014. LSOA-level given for all-age only. GP-level given in five-year age brackets. Number 25 years or over estimated for each LSOA by multiplying the number registered at each GP by the proportion of that GPs register over 25. Estimates calculated for each quarter and averaged across study years.<br>Only consider data for MBCCG GPs and LSOAs within the MBCCG.<br>'Regular' registrations only. 'Temporary' registrations are not counted in the NHS Digital data |
| CDW GP register counts | Y | CDW (primary care records) | Annualised count of the number of adults (25+ years) registered at a MBCCG GP for each LSOA inside the CCG boundaries | Entry date – most recent of study start date (01/04/12), $25^{\text{th}}$ birthday, and GP registration start date.<br>End date – earliest of date of death and GP registration end date proxy. If not relevant then 'NA'.<br>'Regular' registrations only. 'Temporay' registrations not included. |
| Year | Y | NA | Continuous variable form of year | Exploratory analysis suggested a linear trend between (natural logarithm of) GP-registered population and time, hence the use of a continuous form of year. |

| | | | | |
|---|---|---|---|---|
| Missing GPs | Y | NHS Digital | Percentage of the LSOAs GP-registered population missing from the CDW as a result of GPs not in the data sharing agreement | Two GPs not in the data sharing agreement of the CDW. An additional GP closed in September 2015 (before the CDW was created), patients had to register at a new GP so these patients are “missing” pre-September 2015.<br>Percentage calculated using LSOA-level GP registration data released by NHS Digital. We do with calculations with all-age data and assume this variable not to be correlated with age. As with the ’GP-registered population’ variable, mean taken across quarters.<br>For the study years 2012 and 2013, the 2014 value is used. Exploratory analysis suggests this variable does not fluctuate year-on-year. |

Table S2: Description of variables used in the random intercept model of referrals to outpatient respiratory clinics.

| **Variable** | **Time varying (Y/N)** | **Source** | **Description** | **Notes** |
|---|---|---|---|---|
| Outpatient referrals | Y | CDW (secondary care records) | Annualised count of number of referrals to outpatient respiratory clinics | New referrals from GP, for adults aged 25+ years residing within MBCCG boundaries.<br>Clinic inclusion: respiratory, spirometry, lung, or oxygen clinics; nurse or consultant led; at Royal Lancaster Infirmary, Furness General Hospital, or Westmorland General Hospital.<br>Clinic exclusion: post-op, rheumatology, physio, asthma biologics, or sleep clinics, and 2-week-wait cancer referrals. |
| CRD patients | Y | NHS Digital and spatio-temporal model output | Annualised count of number of patients with an asthma, COPD, bronchiectasis, or ILD diagnosis | Patients identified by relevant asthma, COPD, bronchiectasis, and ILD SNOMED CT codes. Additional criterion for asthma diagnosis is an inhaler prescription in the last 12 months. |
| Age | Y | ONS (mid-year estimates) | Percentage of adult (25+ years) population in a given age bracket. | Age brackets '65-74' and '75+' are used; covariate selection methods suggests these are the only relevant age groups. |
| Sex | Y | ONS (mid-year estimates) | Percentage of adult (25+ years) population that are male | |
| COPD prevalence | Y | CDW (primary care records) | Percentage of adult (25+ years) population that have an active COPD diagnosis | Patients identified by relevant COPD SNOMED CT codes. |

| | | | | |
|---|---|---|---|---|
| Distance to hospital | N | OSMR | Travel distance (km) by car to the nearest hospital within the MBCCG | Hospitals considered: Royal Lancaster Infirmary, Furness General Hospital, and Westmorland General Hospital.<br>Distances were calculated using open source routing software in R Studio. Distances were calculated for all 11,594 (as of 14/01/22) postcodes in the study area and then averaged by LSOA. |
| Year | Y | NA | Factor variable form of year | Factor form used as opposed to continuous to better study the evolution of the MBRN in the three years since initiation. |
| MBRN intervention | Y | NHS Digital | Percentage of GP-registered population registered at an MBRN GP | This is calculated for each year regardless of whether the MBRN was yet active in order to account for baseline differences in the areas that have and have not received MBRN intervention.<br>Percentage calculated using LSOA-level GP registration data released by NHS Digital. We do with calculations with all-age data and assume this variable not to be correlated with age. As with the 'GP-registered population' variable, mean taken across quarters.<br>For the study years 2012 and 2013, the 2014 value is used. Exploratory analysis suggests numbers registered at each GP does not fluctuate year-on-year. |

#### 1.3 Demographics summary

Table S3 provides a summary, across all space-time units, of the covariates used in the GLMM for outpatient referrals.

Table S3: Summary of the covariates over all space-time units. All values are percentages, except the distance variable which is in kilometres.

| | **Min** | **1$^{st}$ quartile** | **Median** | **Mean** | **3$^{rd}$ quartile** | **Max** |
|---|---|---|---|---|---|---|
| 65-74 | 4.40 | 13.73 | 16.42 | 16.88 | 20.52 | 32.70 |
| 75+ | 2.33 | 9.90 | 13.54 | 14.09 | 16.79 | 34.71 |
| Male | 40.19 | 46.63 | 47.88 | 48.20 | 49.43 | 65.66 |
| Distance | 0.87 | 3.31 | 6.81 | 9.42 | 13.27 | 38.16 |
| COPD | 0.55 | 2.19 | 2.98 | 3.35 | 4.09 | 10.01 |
| MBRN | 0 | 0.20 | 51.46 | 50.93 | 99.90 | 100.00 |

Table S4 displays the change in mean of the time-varying covariates in the model. Both age variables show a mostly increasing trend indicative of an ageing population, the percentage male variable has increased by a marginal amount, and adult COPD prevalence has consistently increased over the study period. MBRN coverage is unobserved for study years 2012 and 2013 as NHS Digital did not release LSOA-level data for this time frame. Proportion of the population registered at the relevant GPs remained mostly constant between 2014-2017 before increasing in 2018. We assume the 2012 and 2013 values to be equal to 2014.

Table S4: Mean of GLMM covariates by study year

| **Year** | **65-74** | **75+** | **Male** | **COPD** | **MBRN** |
|---|---|---|---|---|---|
| 2012 | 16.0 | 13.5 | 48.0 | 3.16 | - |
| 2013 | 16.5 | 13.7 | 48.0 | 3.24 | - |
| 2014 | 16.8 | 13.9 | 48.0 | 3.30 | 50.7 |
| 2015 | 17.0 | 14.0 | 48.3 | 3.37 | 50.7 |
| 2016 | 17.2 | 14.1 | 48.3 | 3.40 | 50.8 |
| 2017 | 17.3 | 14.2 | 48.3 | 3.44 | 50.8 |
| 2018 | 17.2 | 14.5 | 48.4 | 3.46 | 51.4 |
| 2019 | 17.1 | 14.8 | 48.4 | 3.48 | 51.6 |

Figure S1 illustrates a point made in the main manuscript, that the majority of MBCCG LSOAs have either most or very few of their population registered at a GP that joined the MBRN in 2017. Figure S2 shows the areas in which the MBRN has been active. Lancaster is the only area that has had wide-spread full coverage since all GPs in this area are part of larger, multi-site practices. Although the LSOAs with 0% account for a large amount of the MBCCG spatially speaking, due to the differences in population density (illustrated by the sizes of the LSOAs), phase 1 of the MBRN reached 50% of the total MBCCG population. Note that since the MBRN covariate in the GLMM is time-varying, for the sake of these plots we have used 2017 data only.

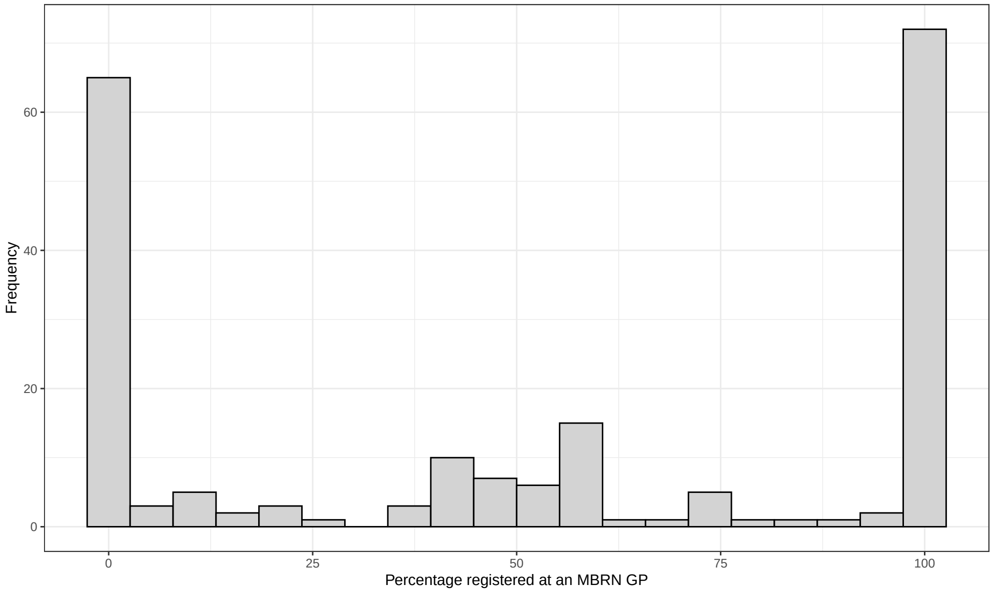


Figure S1: Histogram showing the distribution of the MBRN covariate in the GLMM for study year 2017.

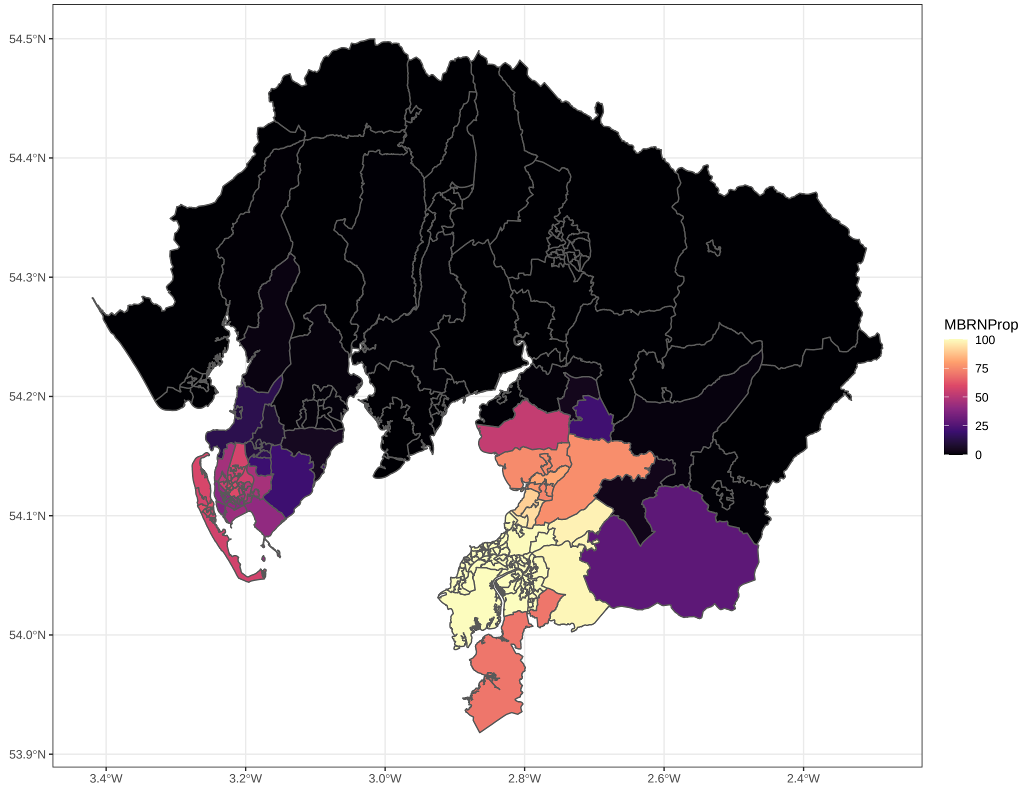


Figure S2: Choropleth map of percentage of LSOA population registered at a GP that joined the MBRN in 2017.

##### 1.4 Outpatient referrals

Figure S3 shows the time trend in the raw referral counts data by intervention area status. For the sake of this figure, we dichotomiose the MBRN covariate so that an LSOA is classed as 'MBRN' if `MBRN` $> 50\%$ and 'Non-MBRN' otherwise. Prior to the initiation of the MBRN, the time trends of the raw counts of referrals are quite similar for both groups, apart from a decrease from areas covered by GPs that joined the MBRN in 2017 (the blue line) between 2012-2013. Post-initiation, the MBRN areas show a dramatic decrease in raw referral count while non-MBRN areas continue in an upward trend. However, the patterns in this plot do not account for population growth or changes in the demographic or health structure of the populations.

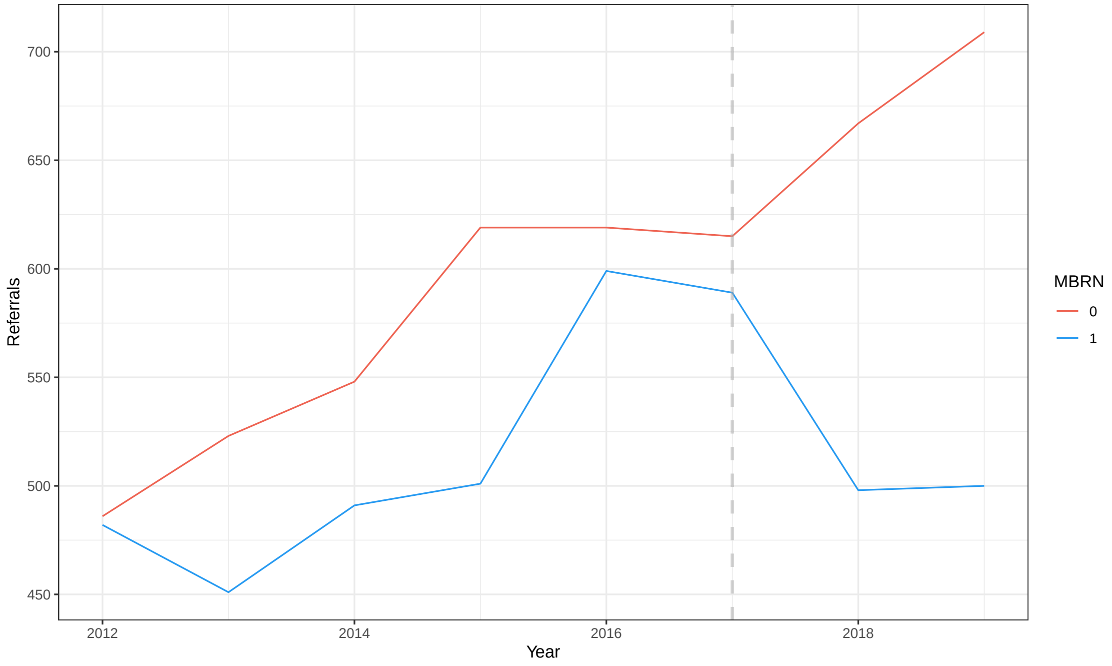


Figure S3: Number of referrals over time for intervention and non-intervention areas. The MBRN covariate has been dichotomised at the 50% mark. The grey dashed line represents the introduction of the MBRN in 2017.

#### 2 Spatio-temporal CAR model methodology

The outcome variable for this model is annualised counts of the number of adults aged 25 years or over registered at a MBCCG GP for each LSOA and study year. Although we are only considering referrals up to March 2020, we utilise data from the year 1$^{st}$ April 2020 to 31$^{st}$ March 2021 for the GP registration model to better study temporal trends and improve prediction capacity. We first consider a generalised linear model (GLM). The outcome counts are sufficiently large (mean = 1181, minimum = 681) to use a log-Gaussian model as an approximation to the Poisson. Covariates are included for (natural logarithm of) CDW estimate (`CDW`), year (`Year`), and proportion of population registered at a GP not in the CDW (`Missing`) which can be calculated from the NHS Digital data.

Let $P_{it}^{NHS}$ be defined as in the main text, then the GLM is of the form:

$$\log(P_{it}^{NHS}) = \beta_0 + \beta_1 \log(\texttt{CDW}_{it}) + \beta_2 \texttt{Year}_t + \beta_3 \texttt{Missing}_{it} \; .$$

The residuals from the GLM exhibit strong spatio-temporal correlation. Spatial autocorreltion was explored using Moran's I statistics computed on the residuals for each year separately; the values ranged from 0.23 to 0.33 with $p$-values less than 0.0001 in all years. The lag-1 temporal autocorrelation calculated for each LSOA separately yielded a mean of 0.3762 across all LSOAs.

Let $S = (S_1, \ldots, S_{T+1})$ denote the set of random effects for time points $t = 1, \ldots, T+1$, where $S_t = (S_{1t}, \ldots, S_{Nt})$ is the vector of random effects for specific time point $t$. Then,

$$\log(P_{it}^{NHS}) \sim \mathrm{N}\left(x_{it}^T \beta + S_{it}, \sigma^2\right) \; .$$

The vector $x_{it}$ denotes the set of explanatory variables, $\beta$ their corresponding regression parameters, and $\sigma^2$ the variance of the residual errors. The model captures the spatio-temporal autocorrelation by assigning the random effects a spatio-temporal extension of conditional autoregressive (CAR) priors, which are a type of Gaussian Markov random field (GMRF). Here we follow the model proposed by Rushworth et al. [1],

$$\begin{aligned} S_t | S_{t+1} &\sim \mathrm{N}\left(\rho_T S_{t+1}, \tau^2 Q(\rho_S, W)^{-1}\right) \qquad t = 1, \ldots, T \\ S_{T+1} &\sim \mathrm{N}\left(0, \tau^2 Q(\rho_S, W)^{-1}\right) \; . \end{aligned} \tag{1}$$

The random effect for time point $T+1$ is specified marginally since $S_{T+2}$ does not exist.

The random effects as specified above are non-separable in space and time. The conditional expectation accommodates for temporal correlation via a first-order autoregressive process with dependency parameter $\rho_T$, where as the spatial autocorrelation is induced via the precision matrix. Numerous specifications for the precision matrix have been made in the CAR literature, but the one used here was proposed by Leroux et al. [2], $Q(\rho_S W) = \rho_S W + (1 - \rho_S) I$ where $\rho_S$ is the spatial dependency parameter, $I$ the $N \times N$ identity matrix, and W an $N \times N$ neighbourhood matrix defined for the 204 non-overlapping spatial units that comprise the lattice data for this study. Using the notation $i \sim j$ to mean "areas $i$ and $j$ share a common border" and $n_i$ to be the total number of neighbours for area $i$, the individual elements of $W$ in the Leroux model are defined as:

$$w_{ij} = \begin{cases} n_i & \text{if } i = j \\ -1 & \text{if } i \sim j \\ 0 & \text{otherwise} . \end{cases}$$

Thus the precision matrix is a weighted average of the spatially dependent and independent structures, and so allows for both weak and strong spatial autocorrelation [3]. The univariate full conditional distribution better illustrates the spatial relationship, and is given by,

$$S_{it} | S_{-it} \sim \mathrm{N}\left(\frac{\rho_S \sum_{j \sim i} S_{jt}}{n_i \rho_S + 1 - \rho_S}, \frac{\tau^2}{n_i \rho_S + 1 - \rho_S}\right) \; ,$$

where $S_{-it}$ is the vector of all random effects at time point $t$ excluding area $i$.

### 3 Modelling decisions for outpatient referrals model

#### 3.1 Measure of deprivation

Table S5 shows the correlation between the covariates for adult COPD prevalence and IMD score as well as the separate domains of the index. There is a strong correlation between COPD prevalence and deprivation quantified by the IMD score, particularly that related to income and employment.

Table S5: Correlation between COPD prevalence and the deprivation scores from the Index of Multiple Deprivation (IMD).

| **Domain** | **Correlation** |
|---|---|
| IMD | 0.740 |
| Income | 0.787 |
| Employment | 0.779 |
| Education | 0.734 |
| Health and Disability | 0.704 |
| Crime | 0.578 |
| Housing and Services | -0.290 |
| Environment | 0.011 |

GLMs were used to compare the use of the IMD score and COPD prevalence covariates, with all other covariates as defined in the model in the main paper. The AIC with the IMD score was 7623 while the AIC with COPD prevalence was 7611.

#### 3.2 Correlation structure

A Poisson GLM model with the covariates outlined in the main article was overdispersed ($\text{mean}(Y_{it}) = 5.5 < 10.0 = \text{var}(Y_{it})$; residual deviance $= 2097 > 1707 = q(0.95, df = 1612)$. Therefore, random effects models were explored.

Moran's I statistic was computed on the residuals of the GLM for each year separately. The statistic for study years 2012-2017 were insignificant. For years 2018 and 2019 the statistics were significant at the 5% level, but only suggested a weak spatial correlation (Moran's I = 0.10 and 0.16 respectively). Therefore, it was concluded that a more complex spatial correlation structure was not necessary and an independent random intercept model was used.

### 4 MCMC Methodology

Samples were drawn from the posterior distributions of the parameters using Markov Chain Monte Carlo (MCMC) methodology.

##### 4.1 Spatio-temporal CAR GP-registration model

The random effects, $S_{it}$ ($i = 1, \ldots, N$ and $t = 1, \ldots, T+1$), act as latent variables in this model and the spatio-temporal CAR prior is as described in (1). For the remaining parameters, the following priors were used:

$$\begin{aligned}
\tau^2 &\sim \text{Inverse-Gamma}\,(0.01, 0.01) \\
\sigma^2 &\sim \text{Inverse-Gamma}\,(0.01, 0.01) \\
\rho_T &\sim \text{Unif}\,(0, 1) \\
\rho_S &\sim \text{Unif}\,(0, 1) \\
\boldsymbol{\beta} &\sim \text{N}\,(\mathbf{0}, 1000\boldsymbol{I}) \ .
\end{aligned}$$

The parameters $(\rho_T, \rho_S)$ were updated via separate random walk Metropolis steps. The tuning parameter was tuned to achieve an acceptance rate between 0.4 and 0.45 in both instances. Alternatively, either of these parameters can be fixed in advance. The parameters $(\tau^2, \sigma^2)$ updated via separate Gibbs sampling steps. The $\boldsymbol{\beta}$ parameters were updated jointly with the latent variables, $\boldsymbol{S}$, using Gaussian Markov random field full conditional sampling techniques outlined in Chapter 2 of Rue and Held (2005).

The independent variables were standardised prior to model fit.

Inference was based on 2,000 independent samples obtained from 250,000 iterations of the algorithm, with the first 50,000 discarded as burn-in and the remaining 200,000 thinned by a factor of 100 to remove any remaining autocorrelation.

##### 4.2 Random intercept outpatient referral model

The random effects, $Z_i$ ($i = 1, \ldots, N$), act as latent variables and have a Normal prior as described in the main article. For the remaining parameters, the following priors were used:

$$\begin{aligned}
\kappa^2 &\sim \text{Gamma}\,(0.01, 0.01) \\
\boldsymbol{\gamma} &\sim \text{N}\,(\mathbf{0}, 1000\boldsymbol{I}) \ .
\end{aligned}$$

The parameter $\kappa^2$ was updated via a Gibbs sampling step. The $\boldsymbol{\gamma}$ parameters were updated jointly with the latent variables, $\boldsymbol{Z}$, using a Gibbs step based on Taylor's second-order expansion approximation techniques outlined in Chapter 4 of Rue and Held (2005).

The independent variables were standardised prior to model fit to reduce multicollinearaity.

Inference was based on 10,000 independent samples obtained from 100,000 iterations of the algorithm, with the first 10,000 discarded as burn-in and the remaining 90,000 thinned by a factor of 9.

#### 5 Results: spatio-temporal CAR GP registration model

Table S6 provides a summary of the outcome measure and covariates used in the GP registration model. Note that the covariate for the year has not been included as this variable simply contains $n$ data points for each of the $T+1$ years in the model.

Table S6: Summary of the covariates used in the GP registration model (NHS Digital GP-registered population, CDW population count, and proportion of the LSOA population registered at a GP not in the CDW) over all space-time units.

| | **Min** | **1$^{st}$** | **Median** | **Mean** | **3$^{rd}$** | **Max** |
|---|---|---|---|---|---|---|
| **NHS Digital** | 680.6 | 960.5 | 1102.5 | 1180.7 | 1356.5 | 2210.7 |
| **CDW** | 365 | 881 | 1048 | 1095 | 1272 | 2153 |
| **Proportion missing** | 0 | 0 | 0 | 0.022 | 0.017 | 0.555 |

Initial, shorter runs of the MCMC algorithm used to fit the spatio-temporal model revealed the temporal dependency parameter, $\rho_T$, to be very close to 1 (0.989), suggesting a very strong temporal autocorrelation in the GP registration data. The model was repeated with $\rho_T$ fixed at 1 which represents perfect temporal auotcorrelation. The Deviance Information Criterion (DIC) is an indicator of model fit for hierarchical models. The DIC for the models with fixed and unfixed temporal temporal dependency were equivalent (DIC=-7299), so for sake of parsimony, we proceeded with the temporal parameter fixed at 1.

Table S7 displays the parameter estimates from the model.

Table S7: Parameter estimates for the spatio-temporal GP registration model.

| **Parameter** | **Coefficient** | **95% CI** | **ESS** |
|---|---|---|---|
| $\rho_s$ | 0.353 | (0.264, 0.450) | 2000 |
| $\sigma^2$ | 0.000173 | (0.000144, 0.000207) | 2138 |
| $\tau^2$ | 0.00263 | (0.00226, 0.00303) | 2000 |
| $\beta_0$ (Intercept) | 6.111 | (6.088, 6.136) | 1360 |
| $\beta_1$ (log(CDW)) | 0.000839 | (0.000817, 0.000860) | 2000 |
| $\beta_2$ (Year) | -0.0152 | (-0.0173, -0.0130) | 1535 |
| $\beta_3$ (Missing GPs) | 1.302 | (1.203, 1.399) | 2000 |

Figure S4 illustrates the spread of the spatio-temporal model prediction of the LSOA-level GP-registered population for study years 2012 and 2013 compared to the observed NHS Digital data for years 2014-2020. The predicted years are very similar to that of 2014. This is supported by census data over the same time period which shows a plateau in the total adult population between 2012-2014, as shown in Figure S5. Although the census population is not identical to the GP-registered population, it is still a good indicator of overall trends.

The predictive performance of the spatio-temporal model was assessed by repeatedly fitting the model on the observed data (study years 2014 to 2020), with one year treated as missing each time

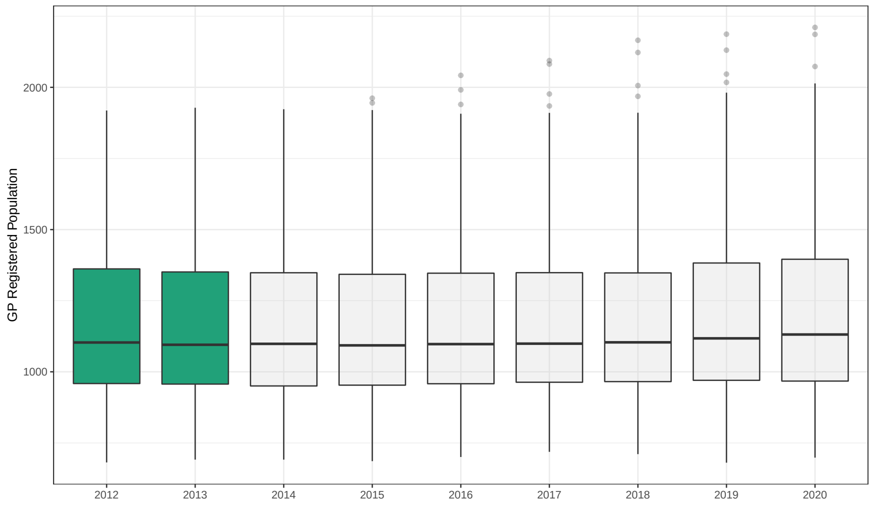


Figure S4: Spread of LSOA GP-registered adult population from NHS Digital data for years 2014-2020 (shown in grey) and spatio-temporal model prediction results for years 2012-2013 (shown in green).

and predicted. The mean absolute percentage error (MAPE) was calculated for the predictions as well as the percentage of LSOA true values within the 95% credible intervals (CI) for the corresponding prediction. The results are displayed in Figure S8. The model predicts well for years 2014-2019, with a maximum of three LSOAs not falling within the 95% CI. The year 2020 performs considerably worse compared to the others, perhaps due to the COVID-19 lockdown affecting normal GP registration behaviour.

Table S8: Mean absolute percentage error (MAE) and proportion of true values that are within the 95% confidence interval (CI) for each year predicted.

| **Year predicted** | **MAPE** | **LSOAs in 95% CI** |
|---|---|---|
| 2014 | 1.81 | 202 (99.0%) |
| 2015 | 1.14 | 202 (99.0%) |
| 2016 | 1.21 | 204 (100.0%) |
| 2017 | 1.02 | 204 (100.0%) |
| 2018 | 1.11 | 201 (98.5%) |
| 2019 | 1.36 | 202 (99.0%) |
| 2020 | 3.28 | 168 (82.4%) |

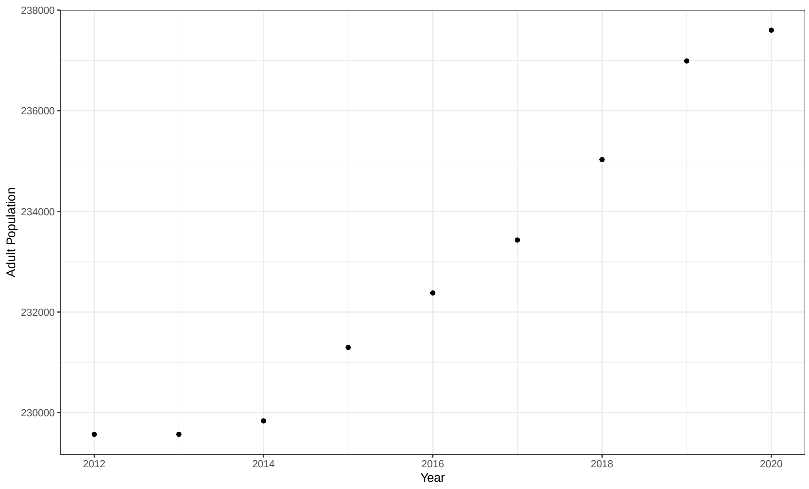


Figure S5: Total population size for the 204 study LSOAs according to ONS mid-year estimates.

### 6 MCMC Diagnostics

Trace plots, density curves, auto-correlation plots, and effective sample size (ESS) calculations were used to assess sufficient mixing of the final chain. Convergence of the MCMC algorithm was established by the Gelman-Rubin convergence diagnostic calculated on three shorter chains to select a suitable length for the burn-in period.

#### 6.1 Spatio-temporal GP-registration model

Figures S6-S8 show the traceplots and density curves for the parameters in the model and a subset of the latent variables. The ESS for the model parameters can be seen in Table S7, whilst the ESS for the latent variables had a median of 2,000 and a minimum of 1,104.

#### 6.2 Outpatient referrals models

Figures S9-S11 show the traceplots and density curves for the parameters in the model. Since there are 21 regression coefficients and 204 latent variables, only a subset of the plots are displayed. The ESS for $\kappa^2$ was 1,239. The ESS for the regression coefficients had a median of 10,000 and a minimum of 8,757, and the latent variables had a median of 10,000 and a minimum of 5,956.

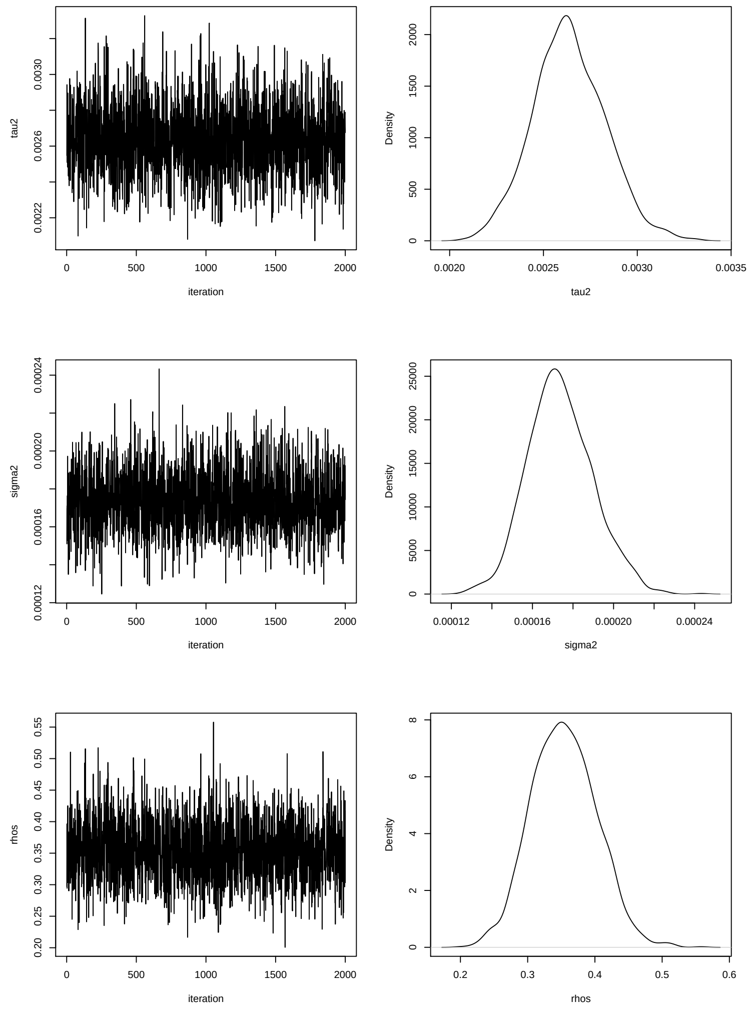


Figure S6: Diagnostic traceplots and density curves for $(\tau^2, \sigma^2, \rho_s)$ in the spatio-temporal GP registration model.

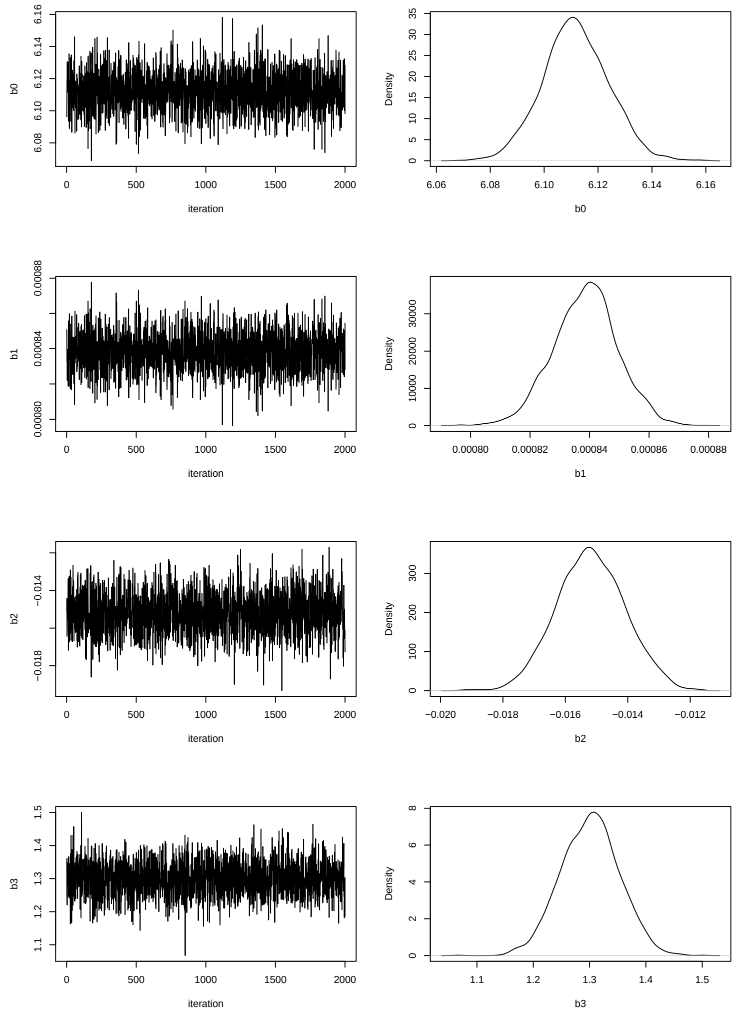


Figure S7: Diagnostic traceplots and density curves for the $\boldsymbol{\beta}$ parameters in the spatio-temporal GP registration model.

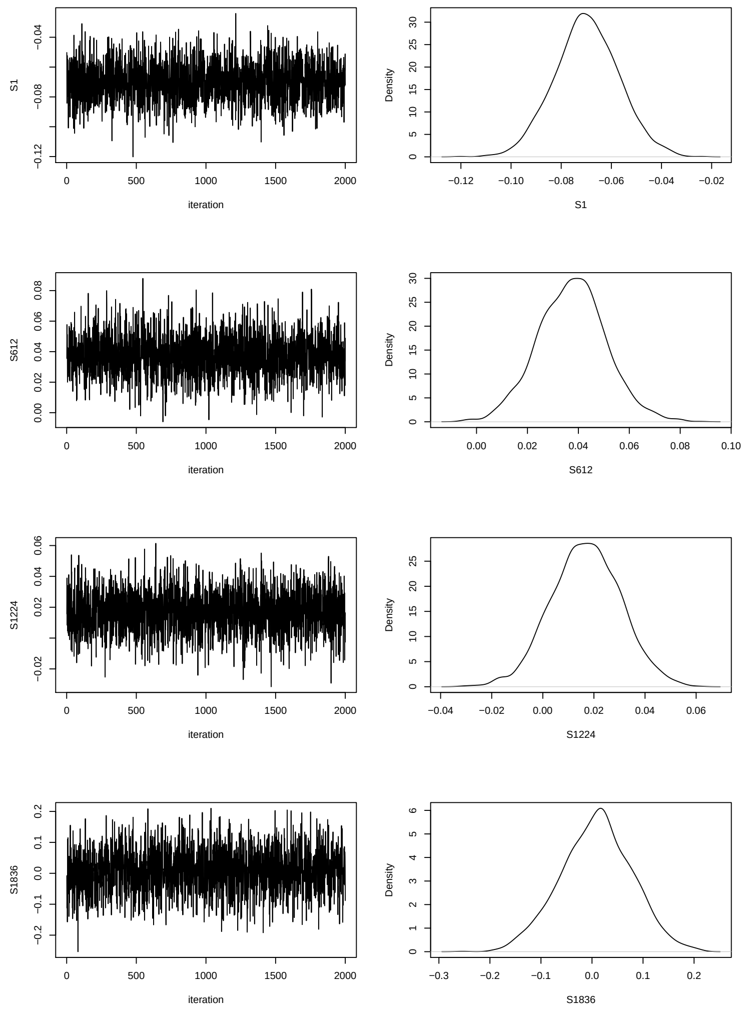


Figure S8: Diagnostic traceplots and density curves for a subset of the $\boldsymbol{S}$ latent variables in the spatio-temporal GP registration model.

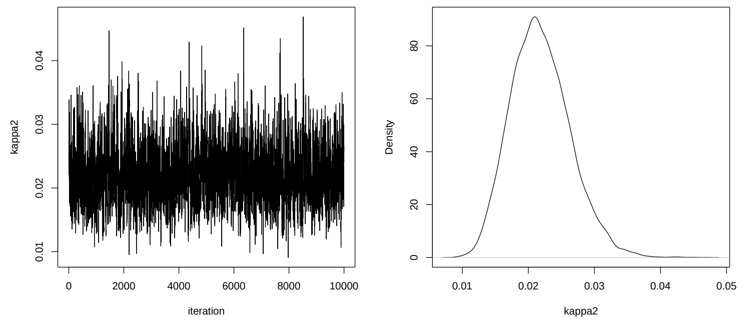


Figure S9: Diagnostic traceplots and density curves for $\kappa^2$ in the random intercept outpatient referrals model.

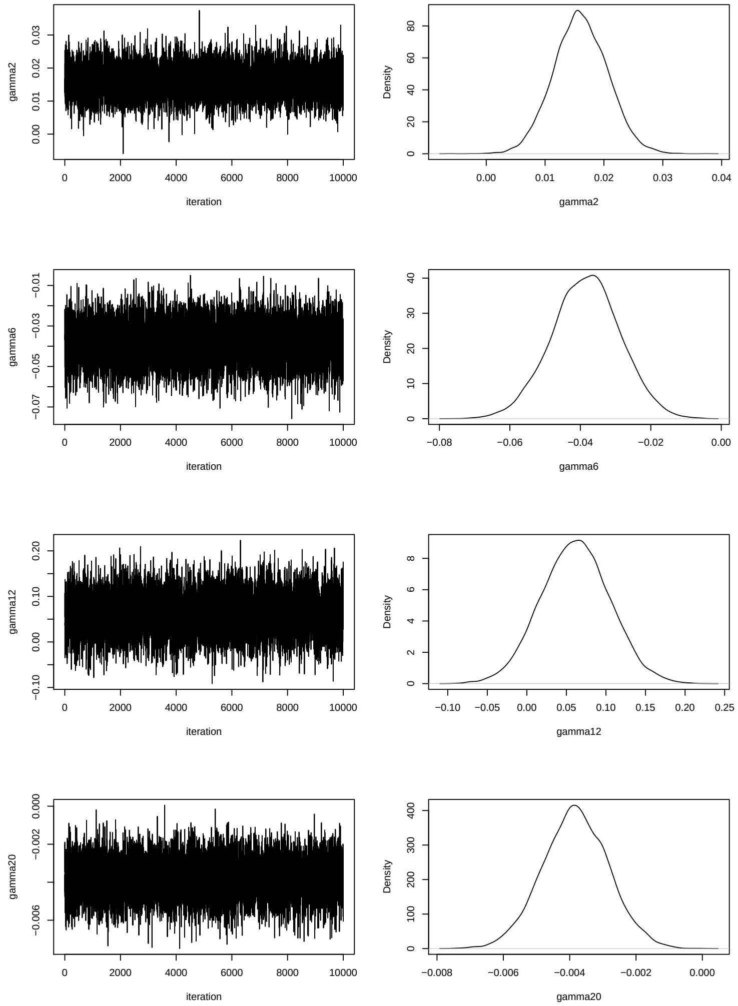


Figure S10: Diagnostic traceplots and density curves for a subset of the regression coefficients, $\boldsymbol{\gamma}$, in the random intercept outpatient referrals model.

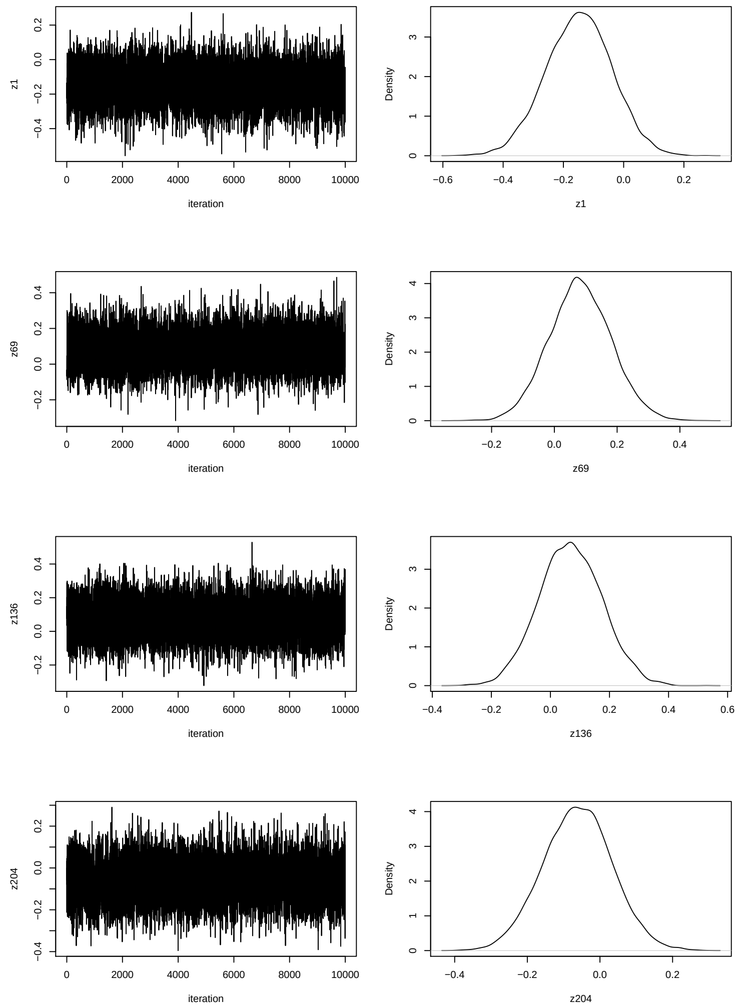


Figure S11: Diagnostic traceplots and density curves for a subset of the $\boldsymbol{Z}$ latent variables in the random intercept outpatient referrals model.
